# Detecting Cardiac Amyloidosis in Italian Cardiology Reports: Structured Variable Extraction versus Direct Free-Text Analysis

**DOI:** 10.64898/2026.01.22.26344604

**Authors:** Sara Mazzucato, Daniele Sartiano, Giuseppe Vergaro, Stefano Dalmiani, Michele Emdin, Silvestro Micera, Calogero Maria Oddo, Claudio Passino, Sara Moccia, Andrea Bandini, Tom Seinen

**Affiliations:** Biorobotics Institute, Department of Excellence in Robotics and AI, Scuola Superiore Sant’Anna, Viale Rinaldo Piaggio 34, Pontedera (Pisa), 56025, Italy; Istituto di Informatica e Telematica, CNR - Consiglio Nazionale delle Ricerche, Via Giuseppe Moruzzi 1, Pisa, 56124, Italy; Department of Medical Informatics, Erasmus University Medical Center, Dr. Molewaterplein 40, Rotterdam, 3015 GD, The Netherlands; Health Science Interdisciplinary Research Center, Viale Rinaldo Piaggio 34, Pontedera, 56025, Italy; Fondazione Toscana Gabriele Monasterio, Via Giuseppe Moruzzi 2, Pisa, 56124, Italy; Bertarelli Foundation Chair in Translational Neural Engineering, Center for Neuroprosthetics and Institute of Bioengineering, Quartier de l’Innovation, Building AIM, Lausanne, 1015, Switzerland; Department of Innovative Technologies in Medicine and Dentistry, Via dei Vestini 31, Chieti, 66100, Italy

**Keywords:** Electronic Health Records, Large Language Models, Natural Language Processing, Named Entity Recognition, Generative AI, Cardiac Amyloidosis, Clinical Decision Support

## Abstract

**Background:** Early and accurate identification of cardiac amyloidosis improves patient outcomes, yet relevant evidence is frequently hidden in free-text records. This study assesses whether structured variable extraction or direct free-text analysis more reliably identifies patients with cardiac amyloidosis, with the goal of informing clinical decision support strategies.

**Methods:** We extracted 21 clinical variables from 432 Italian patient records using supervised and prompt-based methods with both proprietary and locally-deployable computational models. Classification performance was evaluated by comparing extracted data with gold-standard manual annotations. Two feature sets were tested: general clinical variables and amyloidosis-specific risk factors. Additionally, we evaluated direct zero-shot prediction on unstructured clinical notes.

**Results:** For entity extraction, GPT-4.1-mini achieved F1=0.96, comparable to supervised SpaCy (F1=0.95) and GPT-4o (F1=0.94). Local open-source models like Qwen2.5 reached F1=0.94. For cardiac amyloidosis classification, machine learning models using full extracted features matched gold-standard results (SauerkrautLM-Gemma: F1=0.80 vs. gold: F1=0.82). General clinical features alone yielded lower performance (F1=0.68), highlighting that amyloidosis-specific risk factors in unstructured text provide discriminative diagnostic value. Zero-shot direct predictions outperformed supervised feature-based approaches (MedGemma: F1=0.92).

**Conclusions:** Automated extraction and zero-shot prediction effectively structure Italian EHRs and identify amyloidosis patients without manual annotation. Domain-specific risk factors in free-text notes provide substantial predictive value. Italian hospitals can potentially deploy locally-deployable models to screen cardiac amyloidosis without manual annotation or proprietary APIs, enabling privacy-preserving clinical decision support in real-world settings.

## 1 Background

Cardiac amyloidosis (CA) is an infiltrative cardiomyopathy with increasing recognition but persistent diagnostic delays. Early identification improves patient outcomes through timely disease-modifying therapies, yet many patients experience years of misdiagnosis before definitive testing. The diagnostic challenge stems partly from non-specific clinical presentations that overlap with common heart failure etiologies.

Critical diagnostic clues often appear in clinical narratives before formal diagnosis: carpal tunnel syndrome, spontaneous tendon ruptures, spinal canal stenosis, and sensory neuropathies. These red flags remain buried in unstructured free-text anamneses, invisible to structured query-based screening tools [1]. Manual chart review is time-intensive and impractical for systematic screening of large heart failure populations. Automated methods to extract and leverage this information could enable earlier detection and risk stratification. However, most existing NLP tools require expensive manual annotation pipelines or proprietary APIs, limiting adoption in resource-constrained Italian healthcare systems. This study evaluates practical, immediately-deployable approaches using both proprietary and locally-available models, enabling Italian hospitals to structure EHRs without costly annotation workflows or data-sharing requirements.

Electronic Health Records (EHRs) increasingly digitize clinical documentation, combining coded diagnoses with narrative text. While structured fields support research and decision support [2, 3], much diagnostically relevant information remains in free-text notes [4]. Natural Language Processing (NLP) addresses this by converting narratives into structured representations, but most tools target English clinical text [5, 6], limiting applicability in non-English healthcare systems.

Progress in Italian-language clinical NLP has recently accelerated. Recent studies developed transformer-based models for neuropsychiatric entity recognition (F1=0.85) [7], metastasis classification (F1=0.91) [8], prescription appropriateness assessment [9], and stroke variable extraction [10]. Our prior work introduced a structuring method for Italian cardiology EHRs [11].

Large language models enable few-shot and zero-shot learning, reducing reliance on costly annotated datasets [12, 13]. Recent studies explore their use for multilingual clinical tasks [14, 15]. Locally deployable models offer alternatives to proprietary APIs, mitigating privacy concerns [16, 17], though their performance in zero-shot clinical settings requires evaluation.

In this study, we assess the predictive value of unstructured Italian cardiology notes for detecting CA, comparing annotation-based and non-annotation-based approaches for variable extraction. We develop a supervised extraction pipeline using SpaCy and contrast it with prompt-based methods using both proprietary and local models. We evaluate whether extracted variables enable downstream CA classification and examine the feasibility of zero-shot prediction directly from free-text notes.

## 2 Methods

This section provides an overview of the dataset and clinical setting, describes the variables and annotation process, and details the experimental protocol. We present information extraction methods divided into supervised and unsupervised approaches with evaluation metrics, then describe the diagnostic classification stage and evaluation procedures.

### 2.1 Dataset

We used a dataset of 432 heart failure patients collected in clinical practice at Fondazione Toscana Gabriele Monasterio (FTGM) between 2010 and 2024. Among these, 204 were diagnosed with CA through standard diagnostic workup, while 228 had heart failure without amyloidosis. The population was 69.5% male with mean age 75 ± 10 years.

Each patient record included an Italian free-text anamnesis documenting medical history and clinical status at evaluation. These narratives captured cardiovascular risk factors, symptoms, comorbidities, and physical findings as recorded by attending cardiologists. Many records documented amyloidosis-associated conditions (carpal tunnel syndrome, spinal stenosis, neuropathy) that are rarely coded in structured fields but provide diagnostic value for CA screening.

All data were anonymized at source by FTGM staff following GDPR-compliant protocols. The study was approved by the FTGM Ethics Committee (Decree No. 3854, 02/12/2023).

### 2.2 Experimental Setup

Figure 1 illustrates two approaches. First: (i) information extraction using annotation-based (SpaCy) or non-annotation-based (generative models) methods to create feature sets with 12 general features and 21 features including CA risk factors; (ii) classification using machine learning models trained on both sets. Second: zero-shot classification performed directly on unstructured text. This setup compares models trained on manual annotations versus pre-trained models, and evaluates whether classification based on selected features differs from zero-shot approaches leveraging full clinical history.

**Fig. 1.**
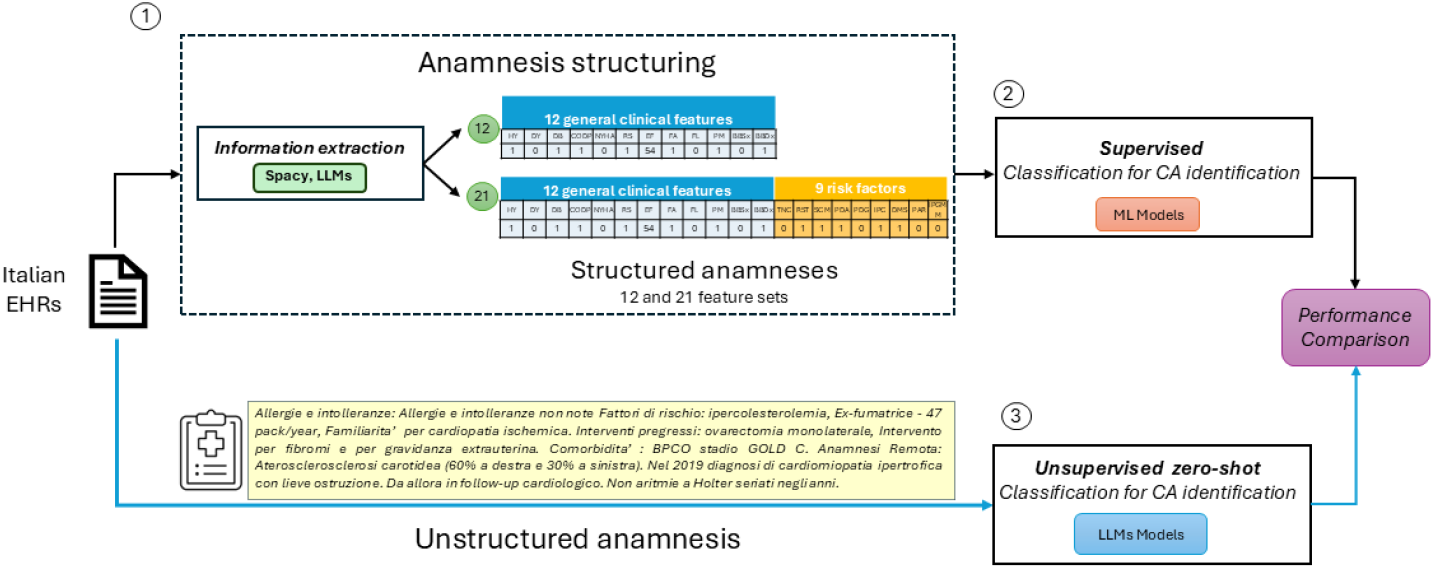
Overview of the study workflow. Italian free-text clinical notes were processed through an extraction pipeline using SpaCy and various LLMs to obtain 12 general features and 9 additional risk factors, forming structured anamneses with 12- and 21-variable sets. These structured features were then used in a supervised pipeline to identify CA. In parallel, zero-shot classification was performed directly on raw clinical texts with LLMs. The workflow is organized into three main steps: (1) information extraction with rule-based and LLM-based approaches; (2) CA classification using supervised models trained on 12 and 21 structured features; (3) zero-shot classification using LLMs directly on unstructured text. Results were compared to evaluate performance and added clinical value.

### 2.3 Variables and Annotation

Twenty-one clinical features were selected based on literature [1] and listed in Table 1. Manual annotation using Label Studio [18] produced a gold-standard dataset. Features included Hypertension (HY), Dyslipidemia (DY), Diabetes (DB), Chronic Obstructive Pulmonary Disease (COPD), New York Heart Association class (NYHA, 1–4), Ejection Fraction (EF, 0–100%), Sinus Rhythm (RS), Atrial Fibrillation (FA), Atrial Flutter (FL), Pacemaker (PM), Left Bundle Branch Block (BBSx), and Right Bundle Branch Block (BBDx).

**Table 1.**
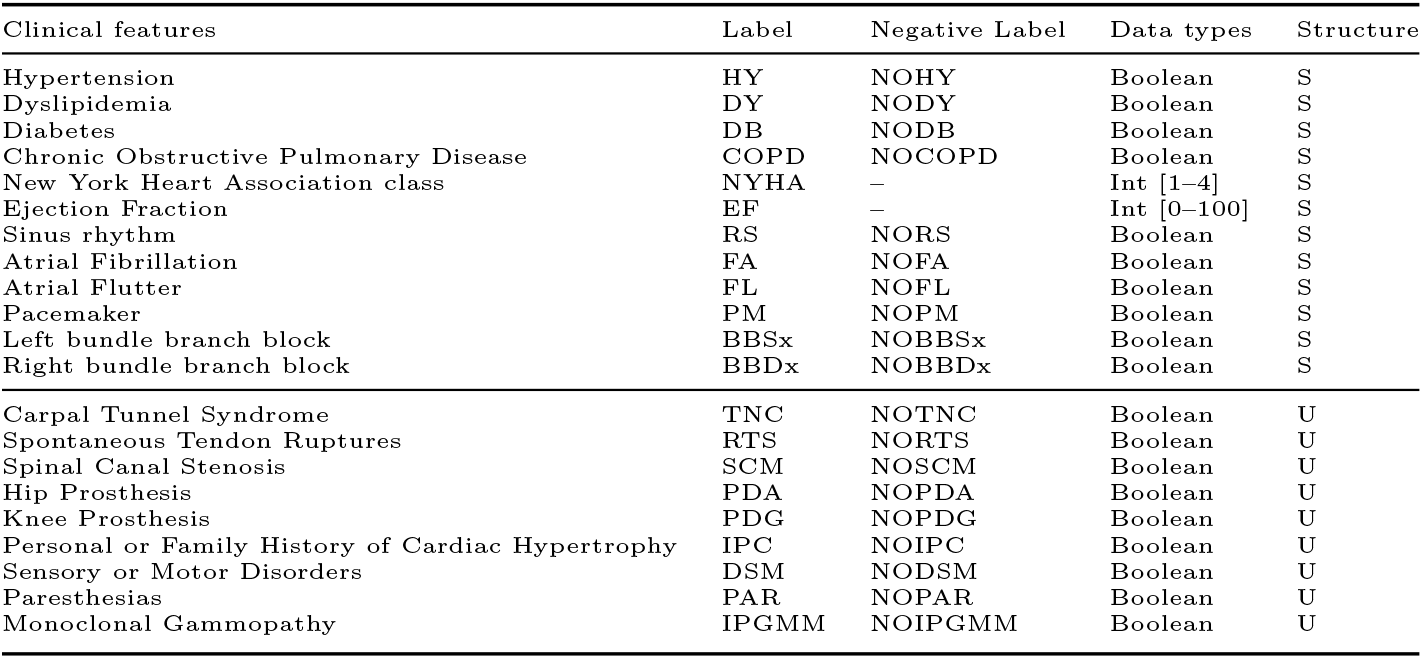
Clinical features of interest extracted from the CA dataset.

| Clinical features | Label | Negative Label | Data types | Structure |
| --- | --- | --- | --- | --- |
| Hypertension | HY | NOHY | Boolean | S |
| Dyslipidemia | DY | NODY | Boolean | S |
| Diabetes | DB | NODB | Boolean | S |
| Chronic Obstructive Pulmonary Disease | COPD | NOCOPD | Boolean | S |
| New York Heart Association class | NYHA | – | Int [1–4] | S |
| Ejection Fraction | EF | – | Int [0–100] | S |
| Sinus rhythm | RS | NORS | Boolean | S |
| Atrial Fibrillation | FA | NOFA | Boolean | S |
| Atrial Flutter | FL | NOFL | Boolean | S |
| Pacemaker | PM | NOPM | Boolean | S |
| Left bundle branch block | BBSx | NOBBSx | Boolean | S |
| Right bundle branch block | BBDx | NOBBDx | Boolean | S |
| Carpal Tunnel Syndrome | TNC | NOTNC | Boolean | U |
| Spontaneous Tendon Ruptures | RTS | NORTS | Boolean | U |
| Spinal Canal Stenosis | SCM | NOSCM | Boolean | U |
| Hip Prosthesis | PDA | NOPDA | Boolean | U |
| Knee Prosthesis | PDG | NOPDG | Boolean | U |
| Personal or Family History of Cardiac Hypertrophy | IPC | NOIPC | Boolean | U |
| Sensory or Motor Disorders | DSM | NODSM | Boolean | U |
| Paresthesias | PAR | NOPAR | Boolean | U |
| Monoclonal Gammopathy | IPGMM | NOIPGMM | Boolean | U |

A second group captured features rarely structured in EHRs: Carpal Tunnel Syndrome (TNC), Spontaneous Tendon Rupture (RTS), Spinal Canal Stenosis (SCM), Hip Prosthesis (PDA), Knee Prosthesis (PDG), Personal or Family History of Cardiac Hypertrophy (IPC), Sensory or Motor Disorders (DSM), Paresthesia (PAR), and Monoclonal Gammopathy (IPGMM). These features add specificity and discriminatory power in evaluating suspected CA patients, highlighting the value of extracting information from narratives to enable comprehensive patient assessment.

### 2.4 Information Extraction

Two approaches were tested: supervised extraction with SpaCy and prompt-based zero-shot extraction using language models.

#### 2.4.1 Supervised Extraction

SpaCy [19] served as a supervised baseline, fine-tuned for token classification on manually annotated clinical notes, achieving F1=0.97 in entity recognition [11]. The model used Italian OPUS and OSCAR corpora [20, 21] with transition-based parsing, Adam optimization, and L2 regularization.

#### 2.4.2 Unsupervised Extraction using Language Models

We employed both proprietary models (GPT-4.1, GPT-4.1-mini, GPT-4o via OpenAI API) and locally-deployable open-source models. Local models included Llama-3 variants (UltraMedical, MMedLlama, OpenBioLLM) [22–24], BioMistral-7B [25], DeepSeek-R1-Distill-Qwen-7B [26], and Italian instruction-tuned models (Medicine-LLM, MedGemma-27B, SauerkrautLM-Gemma-2-9B) [27–29]. Selection prioritized Italian coverage, medical specialization, and deployability.

Models received Italian anamneses and returned JSON objects with predefined variables. The system prompt was: *“You are a medical assistant. Extract structured clinical information from Italian anamnesis text*.*”* The full prompt is in the Appendix.

#### 2.4.3 Evaluation of Information Extraction

The dataset was split 80/20 for training/testing (87 test patients). Both approaches were evaluated using precision (*P*_nlp_), recall (*R*_nlp_), and F1-score (*F* 1_nlp_).

### 2.5 Diagnostic Classification

#### 2.5.1 Supervised Classification

We employed Logistic Regression (LR), Random Forest (RF), and XGBoost (XGB) with grid search optimization. LR provided an interpretable baseline [30]. RF and XGB were tuned for tree depth, sample splitting, learning rate, and regularization to prevent overfitting.

#### 2.5.2 Unsupervised Classification using Language Models

Models performed zero-shot classification on full anamneses, returning binary predictions (1=CA present, 0=absent) and probability estimates (0–1). System prompts requested numeric outputs only, with up to three query attempts to handle parsing errors.

#### 2.5.3 Evaluation of Diagnostic Classification

Datasets structured by language models used 80/20 train-test splits with standardized features and 10-fold cross-validation. SpaCy-structured datasets employed leave-one-out cross-validation. Metrics included accuracy (*A*_cl_), precision (*P*_cl_), recall (*R*_cl_), and F1-score (*F* 1_cl_). Feature importance was assessed using Gini impurity [17], and Spearman correlations evaluated associations between extraction performance and feature importance.

## 3 Results

### 3.1 Descriptive Statistics and Annotation

Figure 2a illustrates binary variable distributions stratified by class, highlighting prevalence differences between CA and non-CA groups. Figures 2b and 2c show EF values and NYHA functional classes across groups.

**Fig. 2.**
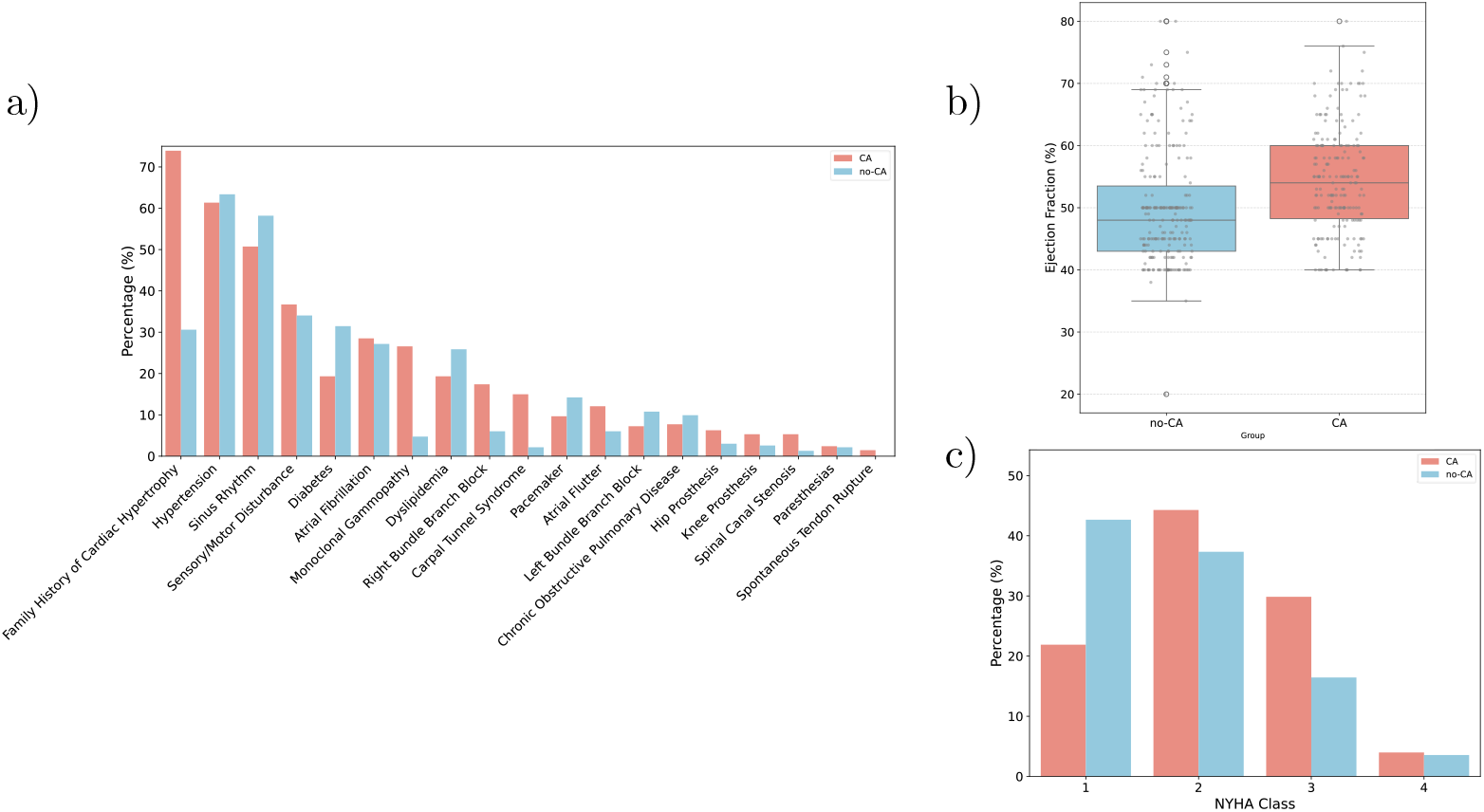
(a) Percentage of positive values in binary variables compared between patients with cardiac amyloidosis (CA) and without (no-CA). (b) Distribution of Ejection Fraction (EF) values, with median, quartiles, outliers, and individual data points. (c) Distribution of New York Heart Association (NYHA) functional classes across the two groups.

### 3.2 Information Extraction

Figure 3a reports the performance of the extraction models. GPT-4.1-mini achieved the highest F1-score (0.96), closely followed by SpaCy (0.95) and other GPT-4 variants. Among open-source models, Qwen2.5-7B-Instruct and MedGemma both exceeded 0.94. SpaCy performed consistently across labels, reaching perfect scores in categories such as ‘RS’, ‘BBDx’, and ‘RTS’. Precision and recall were uniformly high, often near 1.00.

**Fig. 3.**
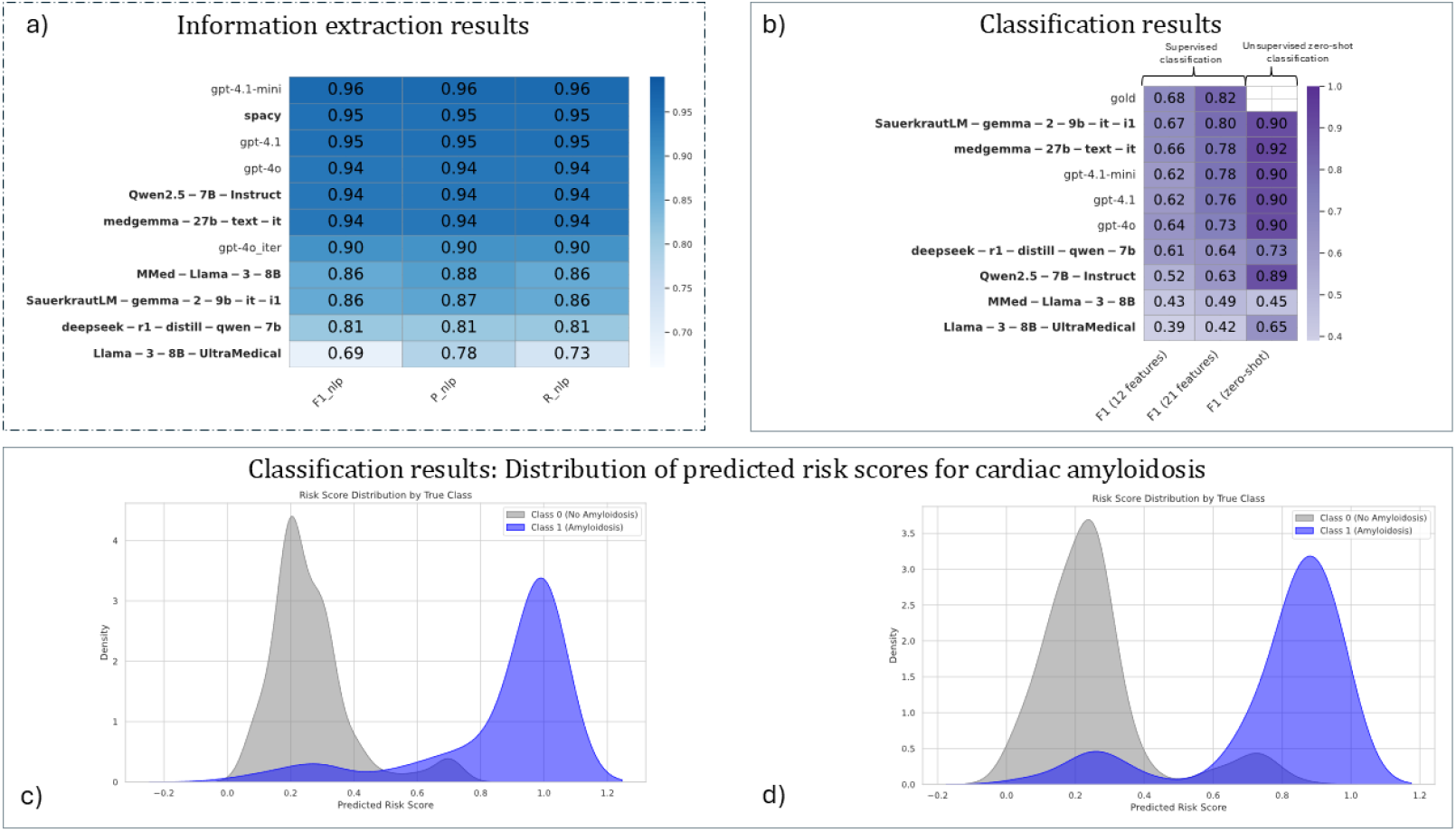
(a) Extraction results: F1, Precision, Recall, and Accuracy, ordered by F1-score with local models in bold. (b) Classification results: F1-scores from the best classifiers using gold annotations, LLM-structured datasets, and zero-shot predictions. Results are shown for 12 features, 21 features, and direct zero-shot classification. (c,d) Distribution of predicted risk scores for CA from the best proprietary model (GPT-4o) and local model (Qwen2.5-7B-Instruct). Blue: CA-positive patients; gray: CA-negative.

GPT-4.1 showed strong performance in ‘HY’ (F1 = 0.94), while Qwen and Llama models exhibited higher variability. For instance, Llama-3-8B-UltraMedical reached 0.88 in ‘COPD’ and 0.96 in ‘BBSx’. SauerkrautLM produced mixed but label-specific results. Iteratively prompting GPT-4o for single features did not improve extraction (F1 = 0.94 vs 0.90).

### 3.3 Classification

Figure 3b compares F1-scores of classifiers trained on annotated and model-generated datasets. With full features, gold annotation classifiers reached F1=0.82, while the best model-based dataset (sauerkrautlm-gemma-2-9b-it-i1) achieved 0.80. Using reduced features, the best score was 0.67. Zero-shot classification with MedGemma-27B-Text-IT peaked at F1=0.92. Detailed results are provided in the supplementary materials.

### 3.4 Relationship Between Extraction and Feature Importance

Figure 4 shows relationships between extraction F1-scores and feature importance from best classifiers. Table E4 reports Spearman correlations. Significant negative correlations (*p <* 0.05) emerged only for SpaCy and MedGemma-27B-Text-IT, suggesting classifiers sometimes assigned higher importance to harder-to-extract but discriminative labels.

**Fig. 4.**
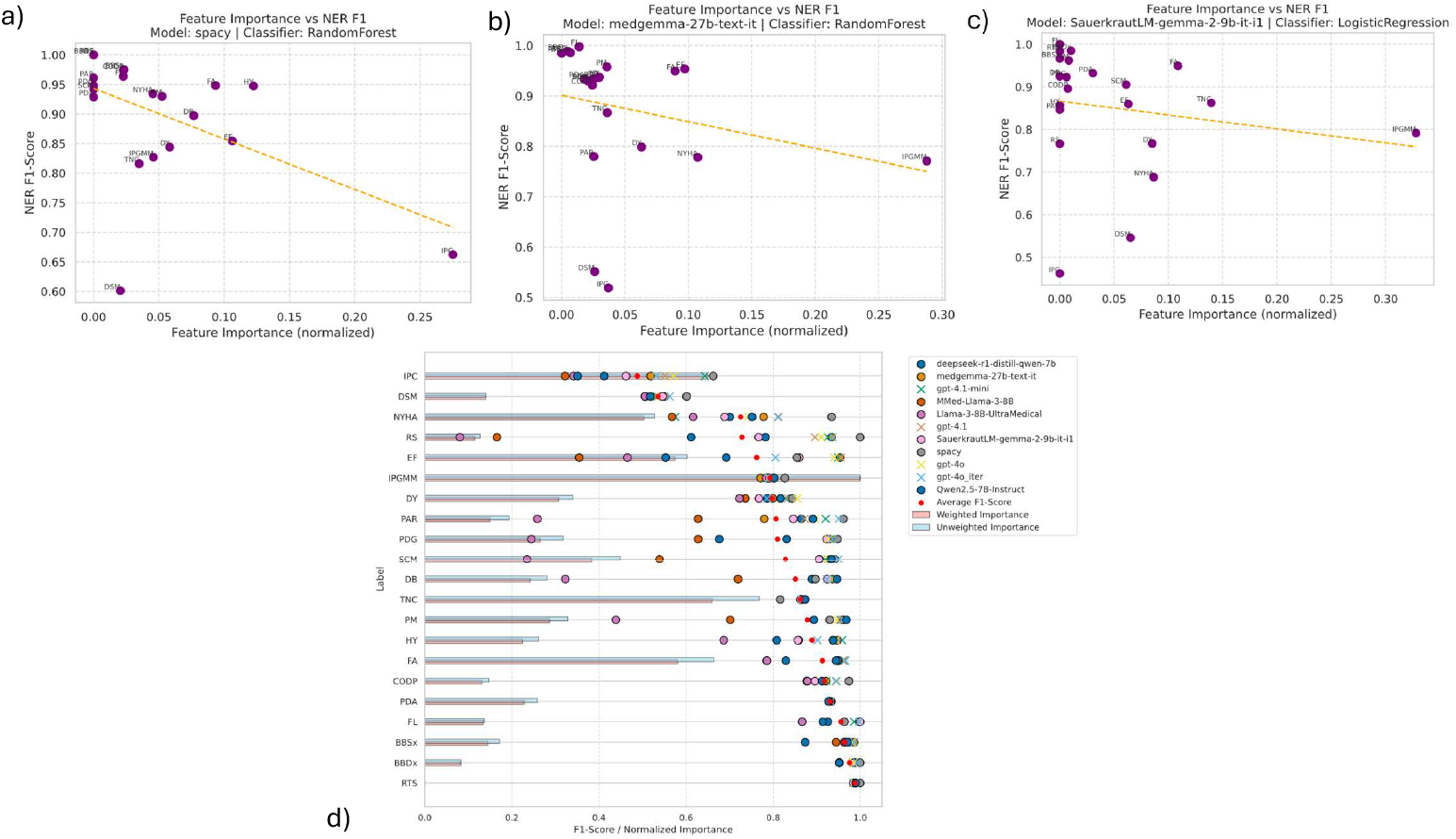
(a), (b), (c) Relationship between feature importance and F1-score per extraction label, evaluated separately for the models. For each model, normalized feature importances (derived from the best-performing supervised classifier trained on 21 structured features) are plotted against the corresponding extraction F1-scores for the same labels. Each point represents a clinical label, with a dashed linear regression line overlaid to highlight the overall trend. This plot aims to explore the alignment between the structural relevance of clinical features and the model’s effectiveness in extracting them from unstructured text. (d) Information extraction performance and feature importance for each clinical label. The scatter points indicate extraction F1-scores achieved by different models, including an average score in red. Markers distinguish between offline and online models. Horizontal bars represent the relative importance of each label as a feature in a downstream classification task, with salmon-colored bars indicating extraction F1-weighted importance and blue bars showing unweighted average importance across the classifiers. Labels are sorted by average extraction performance.

## 4 Discussion

### 4.1 Information Extraction

This study quantitatively evaluated LLMs for structuring Italian EHRs in cardiology, removing the need for costly manual annotation. We assessed proprietary and local transformer-based models on a cardiac anamnesis dataset. GPT-4.1-mini achieved the highest extraction and classification performance, showing that prompt-based LLMs can match or exceed supervised approaches. Strong performance by open-source models highlights opportunities for privacy-preserving, local deployments.

The supervised baseline using SpaCy consistently performed well across most labels. LLMs, including GPT-4 variants, reached F1-scores and accuracy comparable to, or exceeding, SpaCy in some tasks. While SpaCy remained strong in precision and recall, certain LLMs achieved competitive or superior performance, underscoring the importance of selecting models based on application needs.

Iteratively extracting one feature at a time did not improve GPT-4o performance, suggesting the model handles multi-attribute instructions effectively within a single prompt.

### 4.2 Classification

We examined the predictive value of risk factors derived from unstructured notes for CA identification. Using reduced (12 features) and full (21 features) sets, we compared classifiers trained on human-annotated and LLM-annotated data. The classification model using LLM-annotated data nearly matched the model using the manual gold labels, showing that LLM-based annotations can approximate manual ones.

Risk factors extracted from free-text, such as IPGMM, TNC, DSM, SCM, and PDA, ranked among the most important features in the full feature set. Interestingly, some LLMs with modest extraction accuracy still achieved high classification performance. Analysis of feature importance versus extraction F1-score revealed significant negative correlations for SpaCy and MedGemma-27B-Text-IT, suggesting classifiers may rely more on harder-to-extract but discriminative labels.

In zero-shot classification, GPT-4o and Qwen2.5-7B-Instruct achieved the best separation, confirming the robustness of both proprietary and local LLMs for direct CA detection from narratives. Local deployments further support privacy-preserving applications in multilingual or low-resource contexts.

### 4.3 Comparison with Prior Work

Previous studies explored ML-based CA classification using structured data, but few addressed large-scale extraction or text-based classification from Italian notes. Our findings confirm the potential of LLMs for low-resource languages, extending recent advances in transformer-based clinical NLP.

Entity recognition has advanced through rule-based and ML approaches, with SpaCy and other transformers reaching high precision and recall [31]. Our study extends this by evaluating local and general-purpose LLMs for structuring Italian EHRs.

Transformer-based models like BERT set new benchmarks across NLP tasks [32], though most work focused on English. Our results shows similar applicability to Italian texts, in line with GPT-4 findings in few-shot contexts [33].

In related work [11], we applied supervised NER pipelines to Italian cardiology notes, showing structured features could approximate clinician annotations. Here, we broaden the scope by evaluating LLMs for both extraction and zero-shot classification, bypassing manual annotation while reaching comparable performance.

Unlike prior studies focused on English corpora, this work highlights unstructured Italian data and privacy-preserving local deployments, resonating with trends in secure healthcare NLP [34]. Our zero-shot approach contrasts with manual annotation–heavy pipelines [35], offering scalability without major accuracy loss.

While domain-specific models (e.g., BioBERT [36]) often outperform general ones, our results show that prompt-based LLMs can achieve similar outcomes without fine-tuning, echoing findings on clinical BERT adaptations [37].

### 4.4 Limitations and Future Directions

While this study establishes a proof-of-concept that privacy-preserving, locally-deployable NLP models can achieve production-quality performance for Italian cardiology EHRs, limitations include the restricted size and single-center origin of the dataset, which constrains generalizability to other Italian healthcare regions and institutions. However, the controlled setting ensured consistent quality and a solid baseline for future multi-center work. Some entities requiring deeper context, such as IPC and DSM, proved harder to extract, suggesting refinements via prompt engineering, synonym-aware templates, or targeted fine-tuning.

We performed exploratory analysis to identify the relationship between the extraction performance per variable and their feature importance in classification; however further analysis could enhance our understanding. Similarly, deep learning pipelines may reveal additional insights beyond the traditional classifiers used here.

While this study focuses on Italian clinical narratives, the methodological findings are likely generalizable to other non-English healthcare systems. Large language models employed in this work (GPT-4, Llama, Qwen, Gemma variants) are trained on multilingual corpora and show strong performance across diverse languages [14, 15]. Future work should validate the proposed extraction and zero-shot classification approaches on clinical texts in other languages to confirm cross-lingual robustness and establish this as a language-inclusive NLP framework healthcare systems.

The dataset’s limited diversity also restricts broader applicability. Still, ensemble approaches combining SpaCy and LLMs appear promising, merging SpaCy’s fast, deterministic outputs with LLM flexibility [38]. Adding temporal or longitudinal data and expert feedback could further enhance extraction reliability and classifier interpretability [2].

Another important direction for future work is real-world deployment. While the results are encouraging, clinical translation requires addressing several practical challenges. Clinician trust and adoption of AI-generated risk scores have not yet been evaluated in this setting, although evidence from other clinical AI applications suggests that explainability can support acceptance [39]. Integration with existing EHR systems also remains largely unexplored, despite prior successful NLP-based clinical deployments [2]. Finally, cost–benefit trade-offs between models have not been locally validated, even though early analyses indicate that local models may offer viable alternatives [40]. For details on deployment pathways and practical implementation strategies, see the Appendix section.

Despite these challenges, this study shows the feasibility of applying proprietary and local LLMs to Italian narratives for both structured extraction and zero-shot classification. By comparing multiple feature sets and models, and emphasizing privacy-conscious deployments, we outline a practical path toward scalable, interpretable NLP pipelines in healthcare.

## 5 Conclusions

This study shows that automated analysis of Italian cardiology narratives enables effective cardiac amyloidosis screening without manual annotation. Domain-specific risk factors extracted from unstructured text provide substantial diagnostic value beyond routinely coded variables, improving classification performance significantly. Direct analysis of clinical notes outperforms traditional structured variable extraction, suggesting comprehensive narrative review captures diagnostic signals lost in feature reduction. While proprietary models like GPT-4 lead in raw performance, local alternatives offer viable, privacy-conscious options suitable for real-world healthcare applications.

Clinicians and hospital administrators can potentially adopt zero-shot classification with locally-deployable models to screen heart failure populations for cardiac amyloidosis without manual annotation or data-sharing requirements. By bridging the gap between unstructured clinical narratives and automated decision support, this work provides a practical pathway toward scalable, language-inclusive, and privacy-preserving clinical decision support systems accessible to Italian healthcare institutions of all sizes.

## Data Availability

The data that support the findings of this study are not publicly available due to ethical and legal restrictions, but are available from the corresponding author upon reasonable request and with approval from the Ethics Committee.

## List of Abbreviations

Computational and Technical Terms

NLP: Natural Language Processing
LLM: Large Language Models
EHR: Electronic Health Records
NER: Named Entity Recognition
API: Application Programming Interface
JSON: JavaScript Object Notation
GPT: Generative Pre-trained Transformer

Machine Learning Methods

LR: Logistic Regression
RF: Random Forest
XGB: XGBoost
F1: F1-score
P: Precision
R: Recall.

Organizational and Regulatory Terms

FTGM: Fondazione Toscana Gabriele Monasterio
EMC: Erasmus Medical Center
CNR: Consiglio Nazionale delle Ricerche
GDPR: General Data Protection Regulation

## Declarations

### Ethics Approval and Consent to Participate

The study was approved by the FTGM (Fondazione Toscana Gabriele Monasterio) Ethics Committee (Decree No. 3854, 02/12/2023) and conducted in accordance with the Declaration of Helsinki. All participants provided written informed consent prior to inclusion in the study. Clinical records were anonymized at source by FTGM staff prior to data sharing, following GDPR-compliant protocols.

### Consent for Publication

Not applicable. All data presented in this manuscript have been anonymized and do not contain any individually identifiable information.

### Availability of Data and Materials

The datasets generated and/or analysed during the current study are not publicly available due to the protection of patient privacy, but are available from the corresponding author on reasonable request and with appropriate institutional approvals. The code for data processing and analysis is available in the public repository: https://github.com/saramazz/NLP_Italian_EHRs.git, which includes a dedicated CLINICAL IMPLEMENTATION.md guide for practical deployment.

## Competing Interests

The authors declare that they have no competing interests.

## Funding

This work was supported by the “Proximity Care Project” funded by Fondazione Cassa di Risparmio di Lucca. The funder had no role in the study design, data collection, analysis, decision to publish, or preparation of the manuscript.

## Authors’ Contributions

S.M. conceived the study, conducted the analysis, and wrote the manuscript. D.S. contributed to model implementation and evaluation. G.V., S.D., and M.E. provided clinical context and data access. S. Micera, C.M.O., C.P., S. Moccia, A.B., and T.S. supervised the research and provided critical feedback. All authors read and approved the final manuscript.

## Acknowledgements

The authors thank the “Proximity Care Project”, led by Scuola Superiore Sant’Anna di Pisa – Interdisciplinary Center “Health Science” with support from Fondazione Cassa di Risparmio di Lucca, and conducted in collaboration with local healthcare and municipal partners in the Lucca province. Sara also gratefully acknowledges the Department of Medical Informatics at Erasmus Medical Center (EMC) for their support.

## Appendix A Prompt Engineering Details

Each model received a complete Italian anamnesis and was prompted to return a JSON object with a predefined set of clinical variables. The system prompt was: *“You are a medical assistant. Extract structured clinical information from Italian anamnesis text*.*”* The user message included the patient text and specific formatting instructions, as shown below:

`Here is the patient’s anamnesis: {anamnesis}`

Respond with a JSON object like this:

{ “HY”: 1, // Hypertension “DY”: 0, // Dyslipidemia “DB”: 1, // Diabetes “COPD”: “NaN”, // Chronic Obstructive Pulmonary Disease “NYHA”: “II”, // New York Heart Association class “EF”: 55, // Ejection Fraction “RS”: 1, // Sinus Rhythm “FA”: 0, // Atrial Fibrillation “FL”: “NaN”, // Atrial Flutter “PM”: 1, // Pace-maker “BBSx”: 0, // Left Bundle Branch Block “BBDx”: “NaN”, // Right Bundle Branch Block “TNC”: 0, // Carpal Tunnel Syndrome “RTS”: “NaN”, // Spontaneous Tendon Rupture “SCM”: 1, // Spinal Canal Stenosis “PDA”: 0, // Hip Prosthesis “PDG”: 1, // Knee Prosthesis “IPC”: “NaN”, // Personal or Family History of Cardiac Hypertrophy “DSM”: 0, // Sensory or Motor Disorders “PAR”: 1, // Paresthesia “IPGMM”: “NaN” // Monoclonal Gammopathy }

Instructions: – Use 1 if the condition is clearly present – Use 0 if clearly absent – Use “NaN” if not mentioned or unclear – For NYHA, use Roman numerals I–IV or “NaN” – For EF, use a numeric value or “NaN” Only return the JSON object, with no explanation, Markdown, or comments.

## Appendix B Extraction results on the CA dataset

**Table B1.** Extraction results on the CA dataset: Precision (P), Recall (R), and F1-score for Different Models (Part 1)

| Entity | SpaCy |  |  | gpt-4o_iter |  |  | gpt-4o |  |  |
| --- | --- | --- | --- | --- | --- | --- | --- | --- | --- |
|  | F1 | P | R | F1 | P | R | F1 | P | R |
| HY | 94.75 | 94.84 | 94.81 | 90.17 | 90.32 | 90.28 | 94.42 | 94.44 | 94.44 |
| DY | 84.42 | 84.42 | 84.42 | 78.64 | 87.15 | 76.85 | 85.52 | 89.95 | 84.49 |
| DB | 89.72 | 90.32 | 89.61 | 92.37 | 92.38 | 92.36 | 93.65 | 94.08 | 93.52 |
| CODP | 97.40 | 97.40 | 97.40 | 94.41 | 95.20 | 93.98 | 94.41 | 95.20 | 93.98 |
| NYHA | 93.43 | 93.66 | 93.51 | 81.05 | 85.51 | 80.56 | 74.26 | 79.95 | 73.61 |
| EF | 85.44 | 88.92 | 85.71 | 80.53 | 83.44 | 81.02 | 93.98 | 94.02 | 93.98 |
| RS | 100.00 | 100.00 | 100.00 | 93.65 | 97.33 | 90.74 | 91.07 | 96.94 | 86.34 |
| FA | 94.83 | 94.98 | 94.81 | 96.54 | 96.57 | 96.53 | 96.53 | 96.55 | 96.53 |
| FL | 96.37 | 97.17 | 96.10 | 99.77 | 99.77 | 99.77 | 99.77 | 99.77 | 99.77 |
| PM | 93.00 | 95.32 | 92.21 | 95.88 | 95.96 | 95.83 | 94.72 | 95.57 | 94.44 |
| BBSx | 97.53 | 97.92 | 97.40 | 98.61 | 98.61 | 98.61 | 98.09 | 98.09 | 98.15 |
| BBDx | 100.00 | 100.00 | 100.00 | 99.32 | 99.34 | 99.31 | 98.50 | 98.74 | 98.38 |
| TNC | 81.61 | 80.31 | 83.12 | 86.69 | 86.35 | 87.04 | 86.69 | 86.35 | 87.04 |
| RTS | 100.00 | 100.00 | 100.00 | 98.84 | 98.61 | 99.07 | 98.61 | 98.61 | 98.61 |
| SCM | 94.19 | 92.36 | 96.10 | 95.00 | 94.66 | 95.37 | 92.72 | 94.44 | 91.20 |
| PDA | 92.88 | 92.26 | 93.51 | 93.21 | 93.12 | 93.29 | 93.21 | 93.12 | 93.29 |
| PDG | 94.81 | 94.81 | 94.81 | 93.59 | 93.23 | 93.98 | 92.69 | 93.29 | 92.13 |
| IPC | 66.24 | 67.95 | 67.53 | 53.15 | 61.95 | 57.18 | 57.11 | 57.82 | 57.41 |
| DSM | 60.13 | 59.61 | 62.34 | 56.23 | 55.37 | 58.33 | 52.98 | 56.45 | 63.89 |
| PAR | 96.12 | 94.87 | 97.40 | 95.07 | 96.33 | 93.98 | 89.60 | 95.94 | 84.49 |
| IPGMM | 82.69 | 81.54 | 84.42 | 79.16 | 79.95 | 78.47 | 79.05 | 79.68 | 78.47 |
| Overall | 94.75 | 94.84 | 94.81 | 90.17 | 90.32 | 90.28 | 94.42 | 94.44 | 94.44 |

**Table B2.** Extraction results on the CA dataset: Precision (P), Recall (R), and F1-score for Different Models (Part 2)

| Entity | gpt-4.1-mini |  |  | gpt-4.1 |  |  | phi-4-mini |  |  |
| --- | --- | --- | --- | --- | --- | --- | --- | --- | --- |
|  | F1 | P | R | F1 | P | R | F1 | P | R |
| HY | 95.85 | 95.90 | 95.83 | 94.68 | 94.68 | 94.68 | 67.81 | 77.11 | 72.33 |
| DY | 82.83 | 89.14 | 81.48 | 83.44 | 89.10 | 82.18 | 74.55 | 74.11 | 77.21 |
| DB | 92.94 | 93.26 | 92.82 | 93.37 | 93.54 | 93.29 | 79.37 | 82.22 | 78.37 |
| CODP | 92.18 | 91.92 | 92.59 | 94.60 | 95.31 | 94.21 | 81.14 | 89.91 | 76.28 |
| NYHA | 57.51 | 78.95 | 58.80 | 81.23 | 81.22 | 81.25 | 53.10 | 51.81 | 62.33 |
| EF | 94.91 | 94.93 | 94.91 | 95.60 | 95.63 | 95.60 | 51.56 | 67.78 | 57.44 |
| RS | 92.58 | 97.28 | 88.89 | 89.58 | 96.88 | 83.80 | 7.24 | 98.18 | 5.58 |
| FA | 96.31 | 96.40 | 96.30 | 96.07 | 96.07 | 96.06 | 68.85 | 80.74 | 73.49 |
| FL | 98.56 | 98.63 | 98.61 | 99.77 | 99.77 | 99.77 | 96.13 | 96.64 | 96.51 |
| PM | 95.54 | 96.01 | 95.37 | 95.35 | 96.00 | 95.14 | 45.69 | 82.59 | 41.40 |
| BBSx | 97.96 | 98.03 | 97.92 | 97.90 | 97.89 | 97.92 | 90.63 | 92.69 | 89.30 |
| BBDx | 98.50 | 98.74 | 98.38 | 98.30 | 98.63 | 98.15 | 95.49 | 94.04 | 96.98 |
| TNC | 86.69 | 86.35 | 87.04 | 86.69 | 86.35 | 87.04 | 85.61 | 85.89 | 85.35 |
| RTS | 98.38 | 98.61 | 98.15 | 98.49 | 98.61 | 98.38 | 98.25 | 98.60 | 97.91 |
| SCM | 91.88 | 94.63 | 89.58 | 92.56 | 94.71 | 90.74 | 33.96 | 95.78 | 23.49 |
| PDA | 93.21 | 93.12 | 93.29 | 93.21 | 93.12 | 93.29 | 89.50 | 91.86 | 87.44 |
| PDG | 93.12 | 93.70 | 92.59 | 93.40 | 93.78 | 93.06 | 37.25 | 93.39 | 26.28 |
| IPC | 64.28 | 64.60 | 64.35 | 55.13 | 55.58 | 55.32 | 41.71 | 58.28 | 51.86 |
| DSM | 53.39 | 52.50 | 59.95 | 53.06 | 52.15 | 60.19 | 50.96 | 50.57 | 64.19 |
| PAR | 92.08 | 96.36 | 88.66 | 87.26 | 96.12 | 80.56 | 11.09 | 97.73 | 8.14 |
| IPGMM | 78.49 | 79.60 | 77.55 | 79.16 | 79.95 | 78.47 | 81.14 | 82.56 | 85.81 |
| Overall | 95.85 | 95.90 | 95.83 | 94.68 | 94.68 | 94.68 | 67.81 | 77.11 | 72.33 |

**Table B3.**
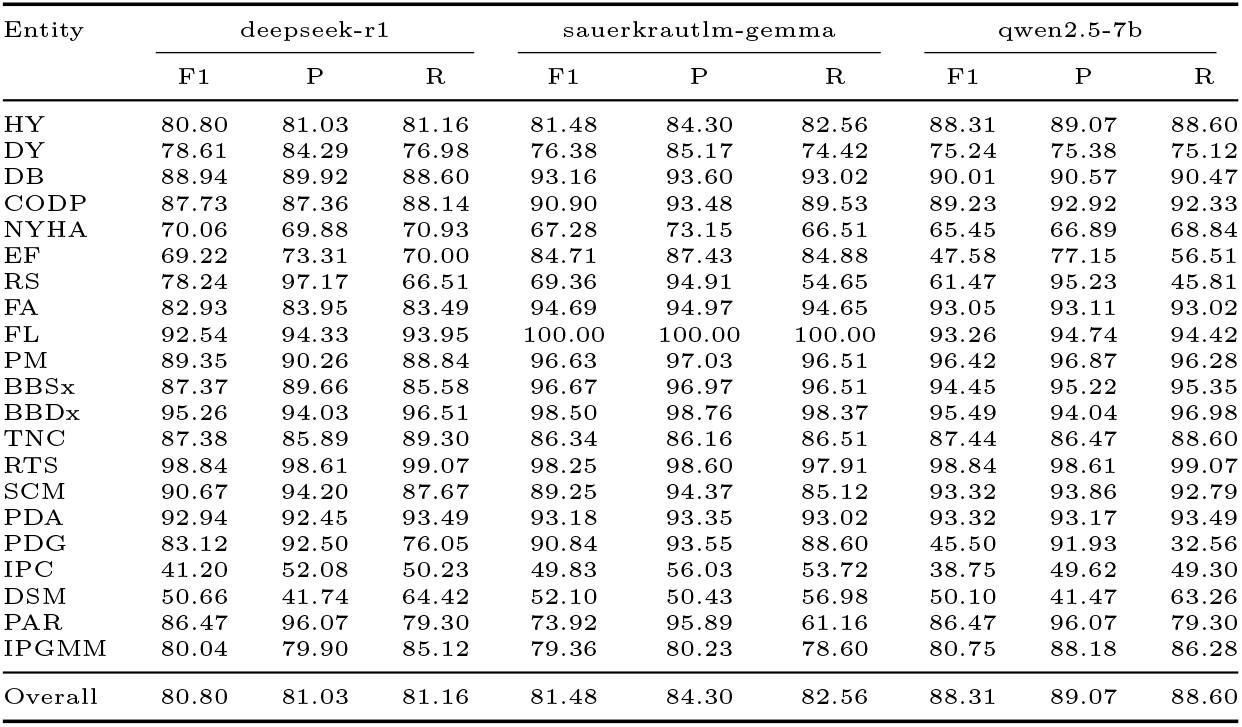
Extraction results on the CA dataset: Precision (P), Recall (R), and F1-score for Different Models (Part 3)

| Entity | deepseek-r1 |  |  | sauerkrautlm-gemma |  |  | qwen2.5-7b |  |  |
| --- | --- | --- | --- | --- | --- | --- | --- | --- | --- |
|  | F1 | P | R | F1 | P | R | F1 | P | R |
| HY | 80.80 | 81.03 | 81.16 | 81.48 | 84.30 | 82.56 | 88.31 | 89.07 | 88.60 |
| DY | 78.61 | 84.29 | 76.98 | 76.38 | 85.17 | 74.42 | 75.24 | 75.38 | 75.12 |
| DB | 88.94 | 89.92 | 88.60 | 93.16 | 93.60 | 93.02 | 90.01 | 90.57 | 90.47 |
| CODP | 87.73 | 87.36 | 88.14 | 90.90 | 93.48 | 89.53 | 89.23 | 92.92 | 92.33 |
| NYHA | 70.06 | 69.88 | 70.93 | 67.28 | 73.15 | 66.51 | 65.45 | 66.89 | 68.84 |
| EF | 69.22 | 73.31 | 70.00 | 84.71 | 87.43 | 84.88 | 47.58 | 77.15 | 56.51 |
| RS | 78.24 | 97.17 | 66.51 | 69.36 | 94.91 | 54.65 | 61.47 | 95.23 | 45.81 |
| FA | 82.93 | 83.95 | 83.49 | 94.69 | 94.97 | 94.65 | 93.05 | 93.11 | 93.02 |
| FL | 92.54 | 94.33 | 93.95 | 100.00 | 100.00 | 100.00 | 93.26 | 94.74 | 94.42 |
| PM | 89.35 | 90.26 | 88.84 | 96.63 | 97.03 | 96.51 | 96.42 | 96.87 | 96.28 |
| BBSx | 87.37 | 89.66 | 85.58 | 96.67 | 96.97 | 96.51 | 94.45 | 95.22 | 95.35 |
| BBDx | 95.26 | 94.03 | 96.51 | 98.50 | 98.76 | 98.37 | 95.49 | 94.04 | 96.98 |
| TNC | 87.38 | 85.89 | 89.30 | 86.34 | 86.16 | 86.51 | 87.44 | 86.47 | 88.60 |
| RTS | 98.84 | 98.61 | 99.07 | 98.25 | 98.60 | 97.91 | 98.84 | 98.61 | 99.07 |
| SCM | 90.67 | 94.20 | 87.67 | 89.25 | 94.37 | 85.12 | 93.32 | 93.86 | 92.79 |
| PDA | 92.94 | 92.45 | 93.49 | 93.18 | 93.35 | 93.02 | 93.32 | 93.17 | 93.49 |
| PDG | 83.12 | 92.50 | 76.05 | 90.84 | 93.55 | 88.60 | 45.50 | 91.93 | 32.56 |
| IPC | 41.20 | 52.08 | 50.23 | 49.83 | 56.03 | 53.72 | 38.75 | 49.62 | 49.30 |
| DSM | 50.66 | 41.74 | 64.42 | 52.10 | 50.43 | 56.98 | 50.10 | 41.47 | 63.26 |
| PAR | 86.47 | 96.07 | 79.30 | 73.92 | 95.89 | 61.16 | 86.47 | 96.07 | 79.30 |
| IPGMM | 80.04 | 79.90 | 85.12 | 79.36 | 80.23 | 78.60 | 80.75 | 88.18 | 86.28 |
| Overall | 80.80 | 81.03 | 81.16 | 81.48 | 84.30 | 82.56 | 88.31 | 89.07 | 88.60 |

## Appendix C Clinical Implementation Guide

For detailed deployment guidance, code examples, and integration workflows, refer to the project repository https://github.com/saramazz/NLP_Italian_EHRs.git. The repository includes a dedicated CLINICAL IMPLEMENTATION.md document describing three practical deployment pathways with cost-benefit analyses and resource requirements.

## Appendix D Classification results on the CA dataset

Confusion matrices, top-10 feature importance rankings, precision–recall curves, and ROC plots illustrate the models’ ability to identify CA patients using the datasets generated by the SauerkrautLM-Gemma-2-9B-IT-i1 classifier, which achieved the highest F1 score for both feature sets.

## Appendix E Statistical analysis

**Fig. D1.**
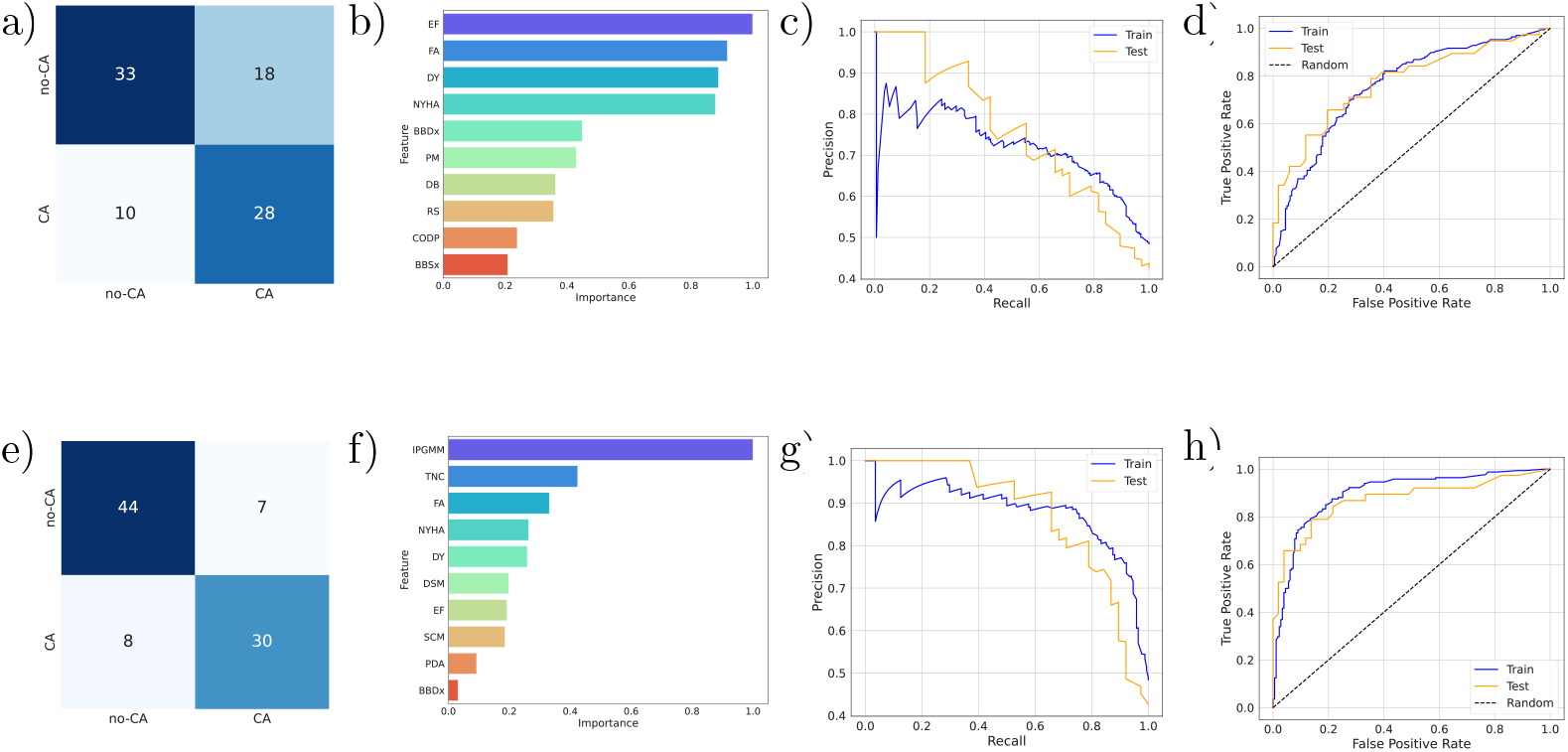
(a,e) Confusion matrix, (b,f) top-10 feature importances, (c,g) precision-recall curve, and (d,h) ROC curve plot for the best-performing classifier (Logistic Regression) trained on the 12-feature set (top) and the 21-feature set (bottom), evaluated on the dataset generated by SauerkrautLM-gemma-2-9b-it-i1 for amyloidosis patient identification.

**Table E4.**
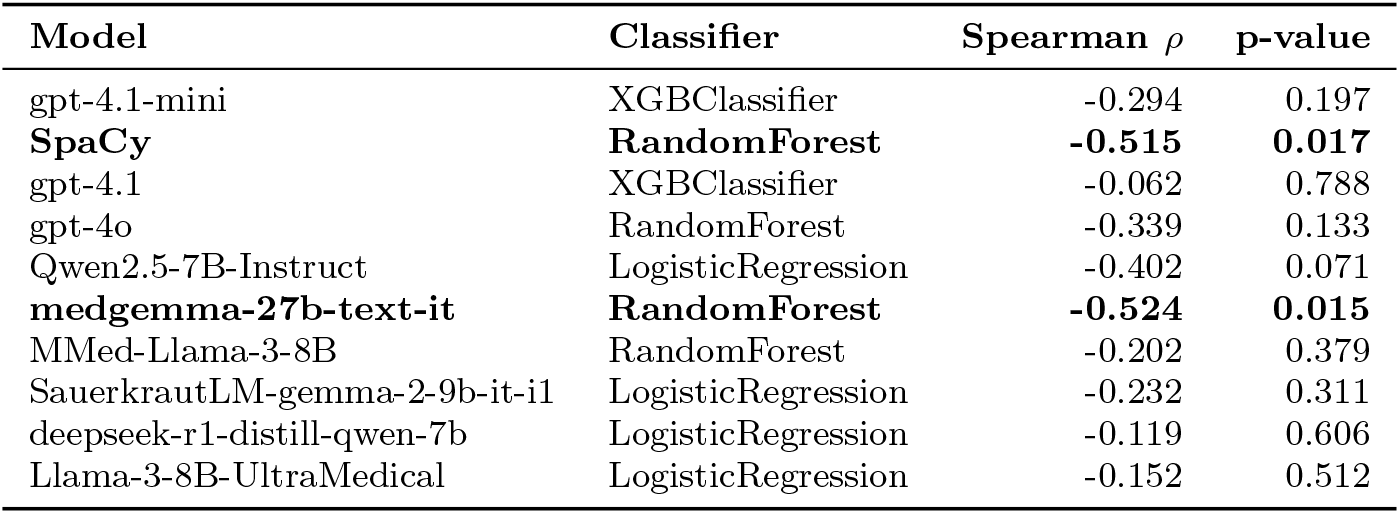
Spearman correlations between normalized feature importances and extraction F1-scores across models and classifiers. Significant correlations (*p* < 0.05) are highlighted in bold.

## Notes

### Competing Interest Statement

The authors have declared no competing interest.

### Funding Statement

This study did not receive any funding

### Author Declarations

Ethics Committee of Fondazione Toscana Gabriele Monasterio gave ethical approval for this work (Decree No. 3854, 02 December 2023)

